# The Physiologic Response to COVID-19 Vaccination

**DOI:** 10.1101/2021.05.03.21256482

**Authors:** Giorgio Quer, Matteo Gadaleta, Jennifer M. Radin, Kristian G. Andersen, Katie Baca-Motes, Edward Ramos, Eric J. Topol, Steven R. Steinhubl

**Author notes:** Correspondence to: Giorgio Quer Scripps Research Translational Institute, 3344 N Torrey Pines Ct Plaza Level, La Jolla CA 92037 USA;. Authors G. Quer and M. Gadaleta equally contributed to the analysis of the data, the discussion, and the preparation of the manuscript.

## Abstract

Two mRNA vaccines and one adenovirus-based vaccine against SARS CoV-2 are currently being distributed at scale in the United States. Objective evidence of a specific individual’s physiologic response to that vaccine are not routinely tracked but may offer insights into the acute immune response and personal and/or vaccine characteristics associated with that. We explored this possibility using a smartphone app-based research platform developed early in the pandemic that enabled volunteers (38,911 individuals between 25 March 2020 and 4 April 2021) to share their smartwatch and activity tracker data, as well as self-report, when appropriate, any symptoms, COVID-19 test results and vaccination dates and type. Of 4,110 individuals who reported at least one mRNA vaccination dose, 3,312 provided adequate resting heart rate data from the peri-vaccine period for analysis. We found changes in resting heart rate with respect to an individual baseline increased the days after vaccination, peaked on day 2, and returned to normal on day 6, with a much stronger effect after second dose with respect to first dose (average changes 1.6 versus 0.5 beats per minute). The changes were more pronounced for individuals who received the Moderna vaccine (on both doses), those who previously tested positive to COVID-19 (on dose 1), and for individuals aged <40 years, after adjusting for possible confounding factors. Taking advantage of continuous passive data from personal sensors could potentially enable the identification of a digital fingerprint of inflammation, which might prove useful as a surrogate for vaccine-induced immune response.

## INTRODUCTION

Due to an unprecedented effort in response to the COVID-19 pandemic, three vaccines are currently authorized and distributed in the United States: two two-dose mRNA vaccines, developed by Pfizer-BioNTech and Moderna, and one single-dose adenovirus-based vaccine, developed by Janssen / Johnson & Johnson.^1-3^ The population-wide efficacy of these vaccines has been well established both through large-scale Phase 3 clinical trials, and reinforced by real-world data.^4-8^ Although it is known that there is substantial variability in individuals’ immune response to vaccines,^9^ and that some fully vaccinated individuals can still become infected,^10^ there is currently no routinely available method to objectively identify a specific person’s response to a vaccine beyond self-reported side-effects, which are common. The Centers for Disease Control and Prevention’s (CDC) V-safe program found a majority (69%) of the 1.9 million enrolled individuals receiving the second dose of a mRNA vaccine reported some systemic side effects.^11^ Many of the reported symptoms were consistent with systemic inflammation including fatigue, myalgias, chills, fever and joint pain being report in the range of 25.6% to 53.9% of individuals the day following their 2^nd^ dose.^12^

In this analysis from the Digital Engagement and Tracking for Early Control and Treatment (DETECT) study,^13^ we collected daily wearable sensor data from the two-weeks before and after each vaccination dose from 4,110 volunteers who documented receiving at least one dose of the vaccine (2,366 received both doses of a mRNA vaccine). We hypothesized that there are digital biomarkers of vaccine-induced inflammatory responses via subtle deviations from an individual’s normal resting heart rate (RHR), as well as changes in a person’s routine sleep and activity behaviors in the days surrounding a vaccine dose. Through exploring individual and vaccine characteristics that might influence that response we identified a stronger response associated with prior COVID-19 infection after the first dose only, and to the Moderna, relative to the Pfizer/BioNTech vaccine, after both doses.

## METHODS

DETECT is an app-based longitudinal prospective study which has enrolled 38,911 individuals so far from the United States (from March 25, 2020 to April 4, 2021) who have donated their wearable data, self-reported symptoms when ill, viral testing results and vaccination dates/type. The protocol for DETECT was reviewed and approved by the Scripps Office for the Protection of Research Subjects (IRB 20–7531). All participants in the study provided informed consent electronically.

Among DETECT participants, 4,110 have reported receiving at least one dose of the vaccine (3,954 first dose only, 2,366 both first and second dose, 156 single dose), 57% were female and their median age was 56 (inter quartile range, IQR 44 - 66).

Individuals who had been vaccinated with the single dose Janssen vaccine were excluded as there were too few (156 individuals) to allow for a meaningful comparison. We have included in the analysis individuals wearing a Fitbit device (76%) and an Apple watch (20%), while 151 individuals with other devices were not included in this analysis. We also excluded 31 participants who reported a vaccine date before Dec. 11, 2020 – the official date of the first US vaccine intake – and 6 participants who did not report age or gender. Individuals were excluded if they had less than 4 days of recording in the 2 weeks before dose 1 vaccination, or less than 3 of the 5 days after vaccination, or less than 14 days during the baseline period (from 60 days to 7 days before vaccination). A number of individuals were excluded in the calculation of RHR (454), sleep (1091) and activity (345) metrics because of missing data.

For each individual, we calculated the average of the absolute changes of RHR, sleep and activity with respect to their individual baseline, which we have previously shown to be relatively stable for an individual over time, but to vary substantially between individuals.^14,15^ A single daily value is considered valid only if the device was worn for more than 15 hours during the day. The RHR was based on the value of heart rate that would be obtained in a supine position immediately after waking but before getting out of bed for Fitbit devices,^14^ and by considering heart rate values over the day by a proprietary algorithm for Apple watches. The individual baseline was calculated using the period from 60 days to 7 days before vaccination, using a decreasing exponential (with exponent α=0.05) to reduce the weight of days farthest in the past. The baseline for the RHR was calculated as

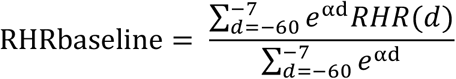

while the RHR metric was

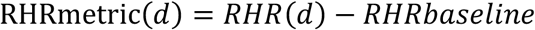

The sleep and activity metrics were calculated accordingly using the total time asleep and the number of steps recorded by the sensor in the 24 hours, respectively. In the figures, the mean (over all individuals) and the 95% confidence interval for each metric (RHR, sleep and activity) in the 15 days before and after the first and second dose of the vaccine are represented. The cumulative distribution of the maximal variation in RHR in the 2 days after the vaccines is also represented.

The cohort of vaccinated individuals (with first and second vaccine doses, treated separately) was then split into subgroups according to gender, age (<40, 40-60, >60), vaccine type received, and if they previously reported a COVID-19 positive test. For each subgroup and for each metric, the mean (over all individuals) of the individual average value (calculated considering the day of vaccination and the following 4 days), with the corresponding confidence interval was calculated. The 95% confidence interval in the calculation of the mean is obtained with a bootstrap method with 1000 iterations.

The demographic characteristics of these groups are reported in the Supplement (Table S.1). Unless stated otherwise, all the reported p-values refer to a two-sided t-test to quantify statistical difference among different groups (Table 1), and to a chi-squared test to evaluate significant changes in the frequency of observation in each group (Table S.1).

**Table 1.** Mean changes in RHR, sleep and activity metrics with respect to the individual baseline in the day of vaccination and the following 4 days, for dose 1 and dose 2. Individuals are divided into binary groups based on gender, age over 60, COVID-19 vaccine type and previously reported COVID-19 positive test.

|  |  | After First Dose |  |  |  | After Second Dose |  |  |  |
| --- | --- | --- | --- | --- | --- | --- | --- | --- | --- |
|  |  | Mean | 95% CI | Individuals | p-value (t-test) | Mean | 95% CI | Individuals | p-value (t-test) |
| RHR Variation [BPM] | Overall | 0.29 | (0.21, 0.37) | 3312 |  | 0.65 | (0.56, 0.76) | 1905 |  |
|  | Female | 0.32 | (0.20, 0.42) | 1893 | 0.44 | 0.7 | (0.56, 0.86) | 1119 | 0.28 |
|  | Male | 0.25 | (0.13, 0.37) | 1419 |  | 0.58 | (0.42, 0.76) | 786 |  |
|  | Moderna | 0.39 | (0.28, 0.51) | 1465 |  | 0.88 | (0.73, 1.03) | 865 |  |
|  | Pfizer/BioNTech | 0.22 | (0.10, 0.33) | 1847 | 0.04 | 0.46 | (0.32, 0.61) | 1040 | < 0.01 |
|  | Age >60 | 0.27 | (0.15, 0.38) | 1348 | 0.66 | 0.53 | (0.40, 0.68) | 957 | 0.03 |
|  | Age <=60 | 0.31 | (0.19, 0.42) | 1964 |  | 0.78 | (0.61, 0.95) | 948 |  |
|  | Prev. Positive | 0.75 | (0.30, 1.20) | 165 |  | 0.28 | (-0.18, 0.73) | 108 |  |
|  | Others | 0.27 | (0.18, 0.35) | 3147 | 0.02 | 0.68 | (0.57, 0.78) | 1797 | 0.1 |
| Sleep Variation [min] | Overall | 2.75 | (1.28, 4.34) | 2675 |  | 5.9 | (3.63, 8.18) | 1490 |  |
|  | Female | 2.51 | (0.41, 4.52) | 1545 | 0.66 | 6.17 | (3.21, 9.06) | 885 | 0.72 |
|  | Male | 3.15 | (0.94, 5.44) | 1130 |  | 5.53 | (2.24, 8.74) | 605 |  |
|  | Moderna | 2.96 | (0.59, 5.18) | 1174 |  | 9.66 | (6.44, 13.33) | 671 |  |
|  | Pfizer/BioNTech | 2.72 | (0.59, 4.75) | 1501 | 0.87 | 2.73 | (-0.26, 5.58) | 819 | < 0.01 |
|  | Age >60 | 2.16 | (-0.19, 4.45) | 1064 | 0.55 | 5.06 | (2.18, 8.02) | 743 | 0.53 |
|  | Age <=60 | 3.19 | (0.95, 5.22) | 1611 |  | 6.52 | (3.56, 9.68) | 747 |  |
|  | Prev. Positive | 7.75 | (0.22, 15.40) | 141 |  | 6.61 | (-3.45, 17.91) | 88 |  |
|  | Others | 2.51 | (0.97, 4.02) | 2534 | 0.15 | 5.81 | (3.59, 8.14) | 1402 | 0.83 |
| Activity Variation [n steps] | Overall | 43.9 | (-24.59, 117.67) | 3421 |  | -363.63 | (-455.50, -278.27) | 1960 |  |
|  | Female | 29.94 | (-60.11, 125.50) | 1971 | 0.69 | -407.18 | (-523.43, -305.41) | 1160 | 0.2 |
|  | Male | 56.97 | (-45.31, 162.52) | 1450 |  | -295.73 | (-430.40, -147.68) | 800 |  |
|  | Moderna | -4.61 | (-106.04, 93.00) | 1516 |  | -583.83 | (-712.91, -457.53) | 880 |  |
|  | Pfizer/BioNTech | 81.07 | (-15.57, 182.10) | 1905 | 0.23 | -185.04 | (-312.38, -61.69) | 1080 | < 0.01 |
|  | Age >60 | -80.71 | (-177.65, 20.72) | 1375 | < 0.01 | -337.26 | (-453.95, -227.53) | 969 | 0.59 |
|  | Age <=60 | 126.82 | (28.46, 223.58) | 2046 |  | -392.36 | (-525.60, -261.86) | 991 |  |
|  | Prev. Positive | -190.16 | (-435.46, 55.30) | 176 |  | -192.74 | (-560.89, 152.98) | 117 |  |
|  | Others | 55.84 | (-18.38, 127.25) | 3245 | 0.13 | -374.93 | (-455.95, -289.25) | 1843 | 0.37 |

A multiple linear regression model was used to calculate the estimated marginal means after adjusting for potential confounding variables. A t-test was used to assess significant coefficients and the associated p-values (Table S.2).

## RESULTS

At least one vaccination to date was reported by 4,110 participants in the DETECT study. After applying the exclusion criteria discussed in Methods, we included a total of 3,312 (80%) individuals for the analysis of changes in their RHR. Of them, 165 (5.0%) reported having been previously diagnosed with COVID-19 infection, 1,465 (44%) received the Moderna vaccine and 1,847 (56%) received the Pfizer-BioNTech vaccine. 3,421 (83%) and 2,675 (65%) participants contributed adequate data - as discussed in Methods - to evaluate changes in activity and sleep, respectively. (Table S.1)

We observed that the average RHR significantly increased the day following vaccination, reaching a peak on day 2 with a population mean increase of +0.49 (CI: [0.38, 0.61], one-sided t-test, p = 0.012) and +1.59 (CI: [1.40, 1.74], p < 0.01) beats per minute (BPM) with respect to baseline, following the first and second dose, respectively. We found that the average RHR did not return to baseline until day 4 after the first dose and day 6 after the second (Figure 1a & b) The majority of vaccinated individuals, 70% and 76% after first and second dose, respectively, experienced an increase in their RHR in the two days following the vaccine. (Figure 1c & d) We explored several participant and vaccine characteristics that could impact immune response. (Table 1) We found that average RHR changes with respect to baseline in the 5 days following vaccination did not vary by gender (two-sided t-test, p = 0.44 and p = 0.28 for first and second doses, respectively). In contrast, we found that RHR responses vary by age, with individuals age < 40 years having the greatest increase in RHR. (Figure 2) We showed that < 60 years was associated with a significantly higher RHR increase than 60+ years, but only after the second dose of the vaccine (average 0.78 versus 0.53 BPM, p = 0.03).

**Figure 1.**
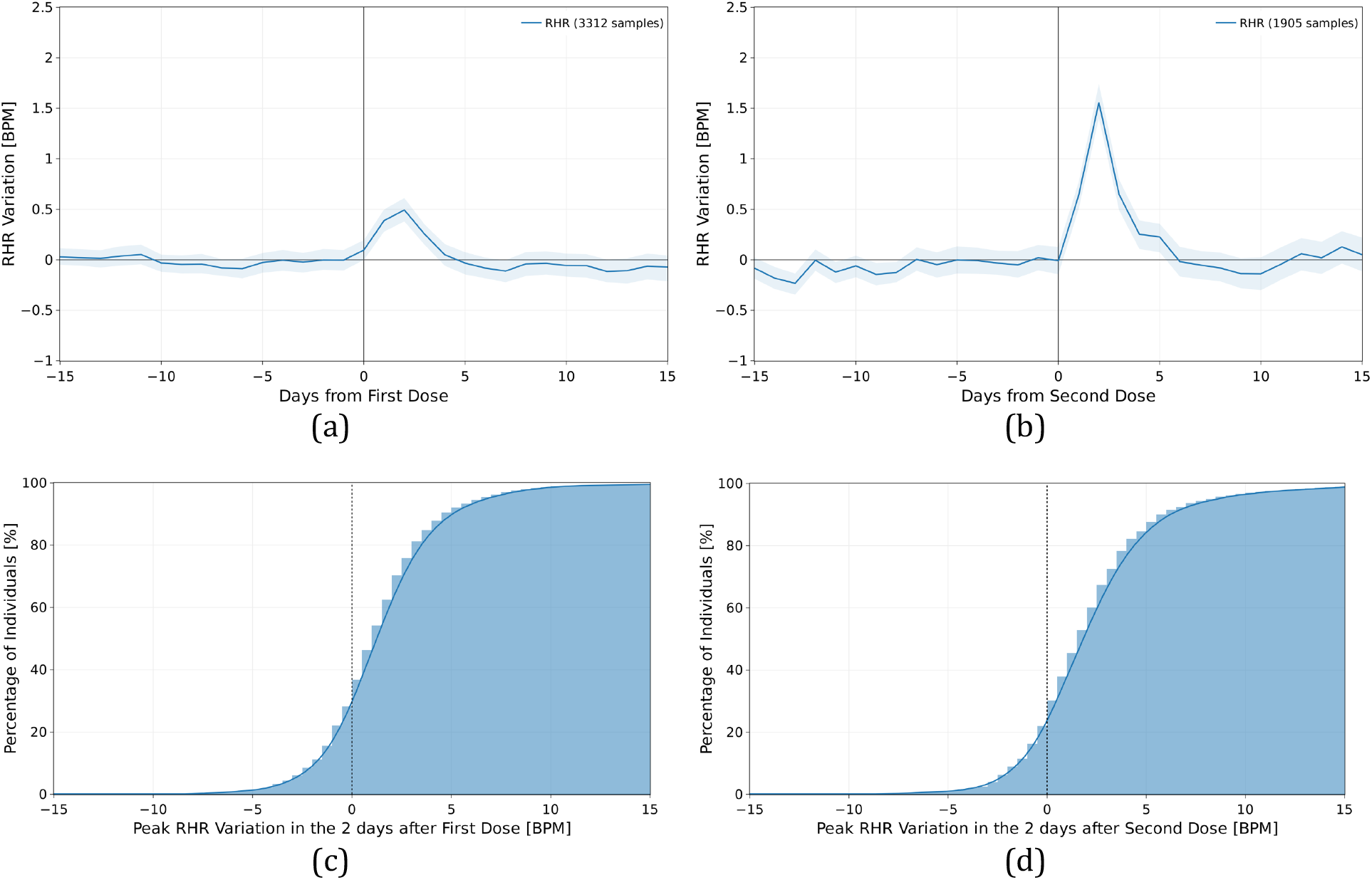
Mean and 95% confidence interval of the absolute individual changes in resting heart rate (in BPM) with respect to the individual baseline around the date of vaccination (day 0), for the first dose of the vaccine (a) and for the second dose (b). The cumulative distribution of the maximal variation in resting heart rate in the 2 days after the vaccines after the first (c) and second (d) vaccine dose.

**Figure 2.**
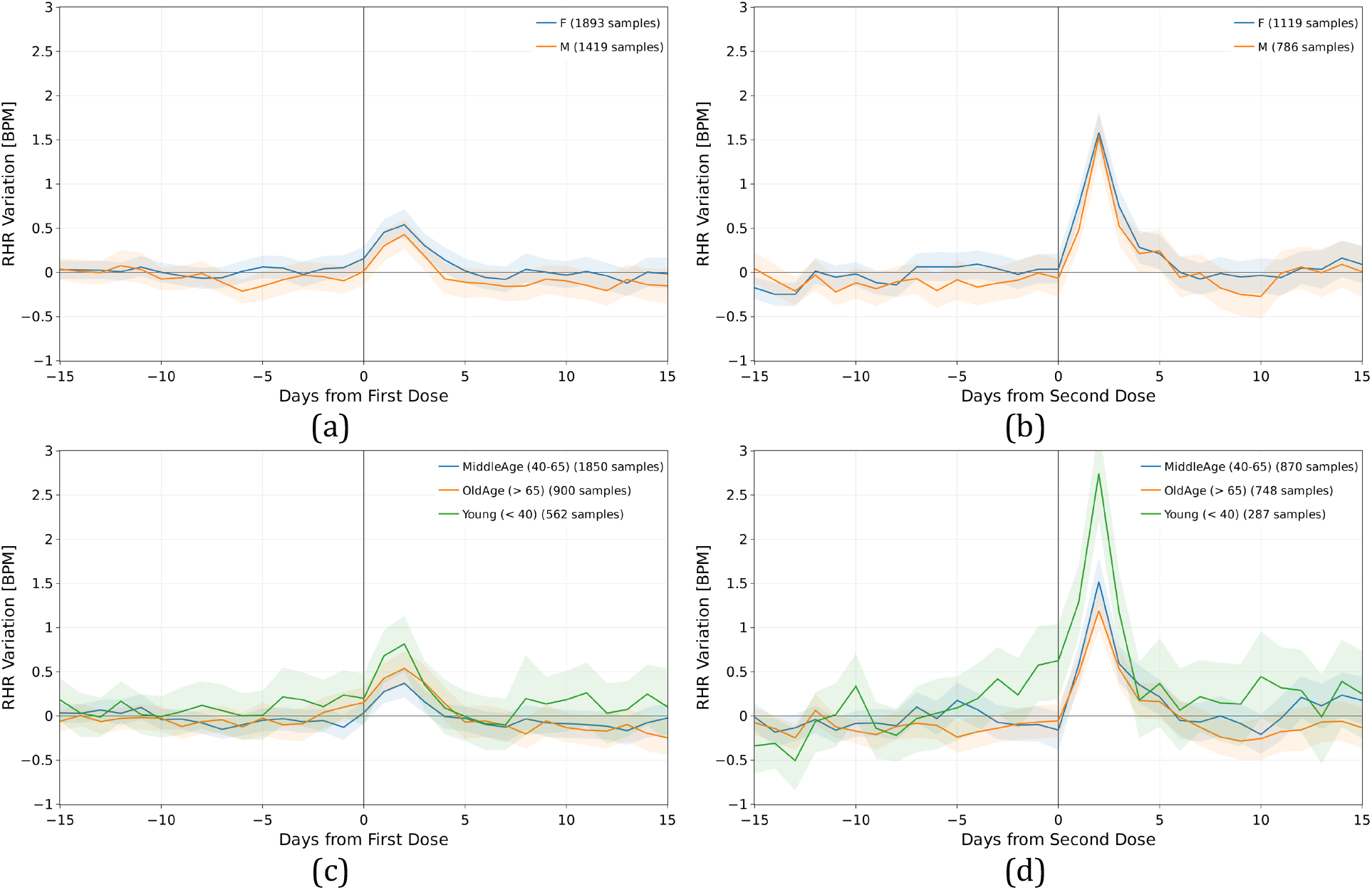
Mean and 95% confidence interval of the absolute individual changes in resting heart rate (in BPM) with respect to the individual baseline around the date of vaccination (day 0), for the first dose of the vaccine (a), (c), and for the second dose (b), (d), for all individuals grouped by gender (a), and (b) and age(c) and (d).

We found that prior COVID-19 infection was associated with a significantly higher RHR increase after the first vaccine dose relative to those without prior infection (average 0.75 versus 0.22 BPM, p = 0.02), but no significant difference after the second dose (0.28 versus 0.68, p = 0.10). (Figure 3a & 3b)(Table 1) The changes in RHR for individuals who received the Moderna vaccine were significantly greater than those who received the Pfizer-BioNTech vaccine, after both the first (0.39 versus 0.22, p = 0.04) and second doses (0.88 versus 0.46, p < 0.01). (Figure 3c & 3d)(Table 1)

**Figure 3.**
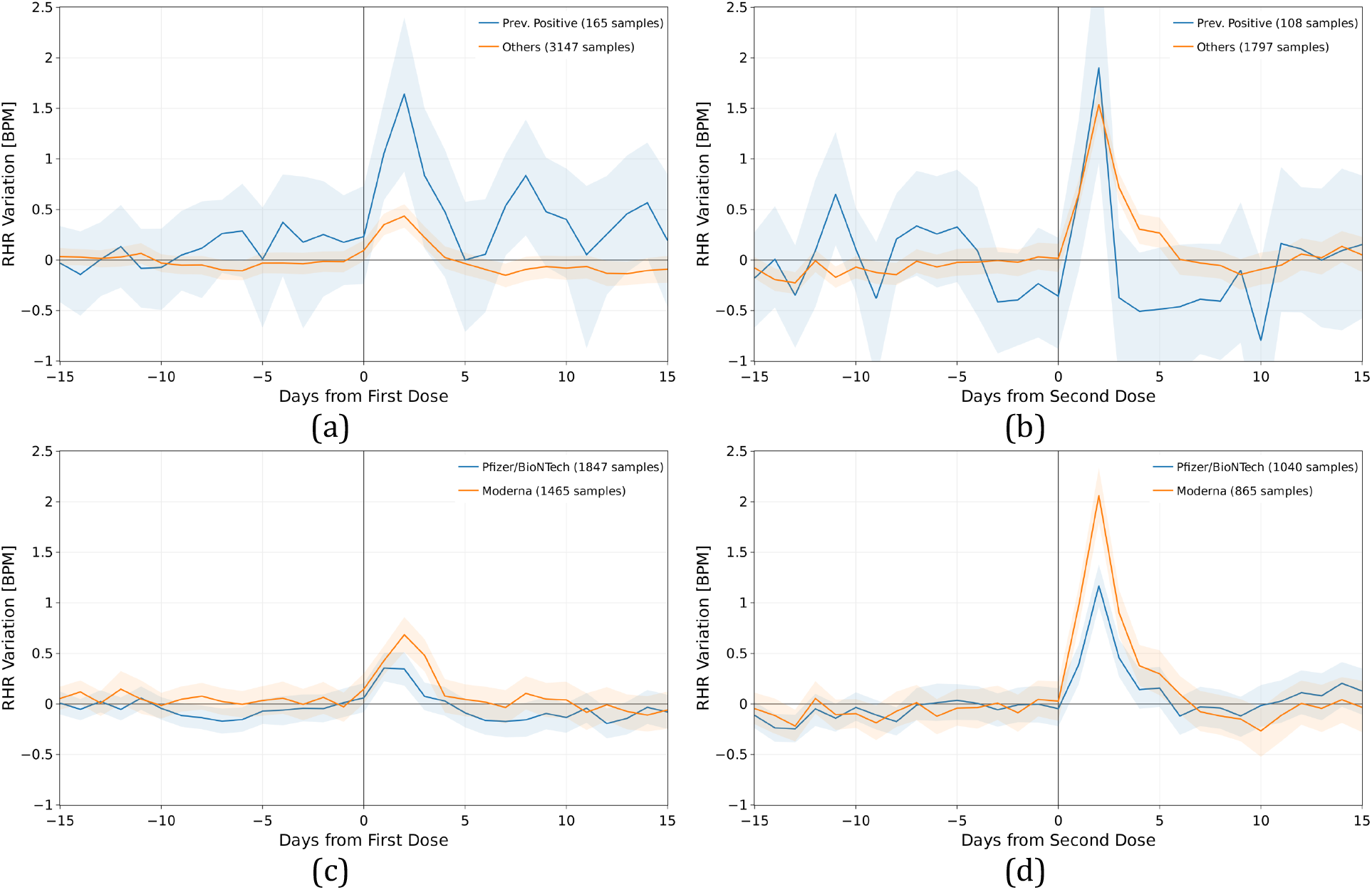
Mean and 95% confidence interval of the absolute individual changes in resting heart rate (in BPM) with respect to the individual baseline around the date of the first and second dose of vaccine (day 0), based on prior COVID-19 infection (a) and (b), and based on the type of mRNA vaccine received, either Pfizer-BioNTech or Moderna vaccines (c) and (d).

A multiple regression model was used to adjust for potential confounding factors. Prior COVID-19 infection was independently associated with a higher RHR increase after the first dose, with estimated marginal mean of 0.770 (CI: [0.392, 1.147]) versus 0.293 (CI: [0.195, 0.392]) BPM, p=0.014; and no significant difference after the second dose, 0.275 (CI: [-0.183, 0.734]) versus 0.731 (CI: [0.606, 0.856]), p=0.055, after adjusting for age, gender, device, and vaccine type. Similarly, the Moderna vaccine was also independently associated with a higher RHR increase after both doses (with respect to Pfizer-BioNTech), with estimated marginal mean of 0.622 (CI: [0.400, 0.843]) versus 0.442 (CI: [0.231, 0.652]), p=0.035 for first dose, and 0.721 (CI: [0.453, 0.989]) versus 0.285 (CI: [0.023, 0.548]), p<0.01 for second dose, after adjusting for age, gender, device, and prior COVID-19 infection. Age was also associated with RHR response, but only after the second vaccine dose (p<0.01). We assessed the interaction between age and gender but did not find it to be significant (p=0.53 and p=0.52 for first and second dose, respectively). (Table S.2)

We also observed that normal activity and sleep patterns among participants were minimally affected by the first dose of the vaccine, with no decrease in number of steps and a mean increase of only 13 minutes (CI: [10, 16]) of sleep in the day following the vaccine. However, a significant decrease in activity (−1375 steps, CI: [-1540, -1205]) and increase in sleep (40 minutes, CI: [34, 45]) relative to baseline were observed on day 1 after the second vaccine dose, both of which returned to baseline by day 2. (Figures 4 and 5)

**Figure 4.**
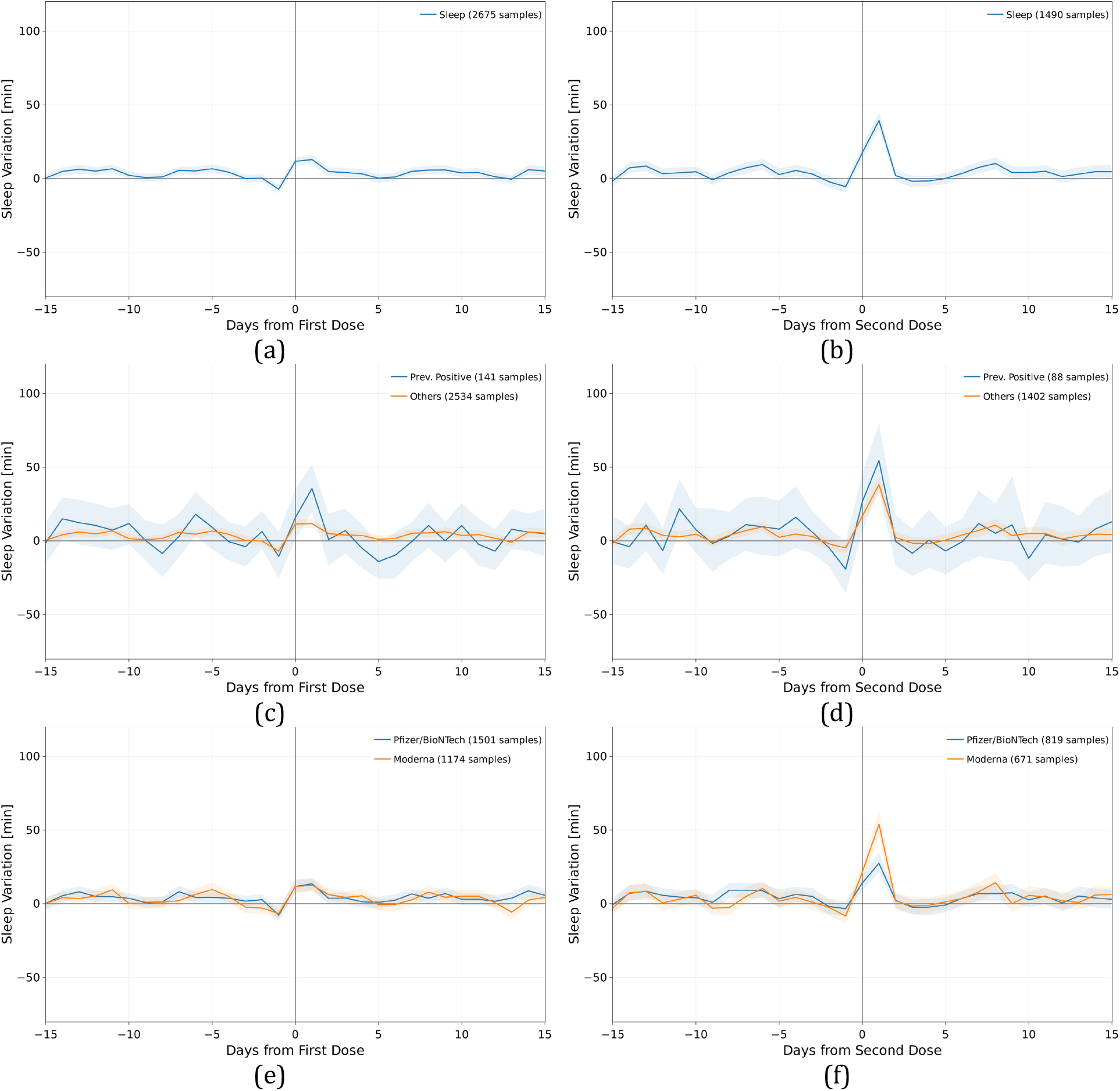
Mean and 95% confidence interval of the absolute individual changes in sleep metric (in minutes) with respect to the individual baseline around the date of vaccination (day 0), for the first dose of the vaccine (a), (c), (e), and for the second dose (b), (d), (f), for all individuals vaccinated (a) and (b), for individuals previously tested positive to COVID-19 (c) and (d), and for individuals vaccinated with the Pfizer-BioNTech or Moderna vaccines (e) and (f).

**Figure 5.**
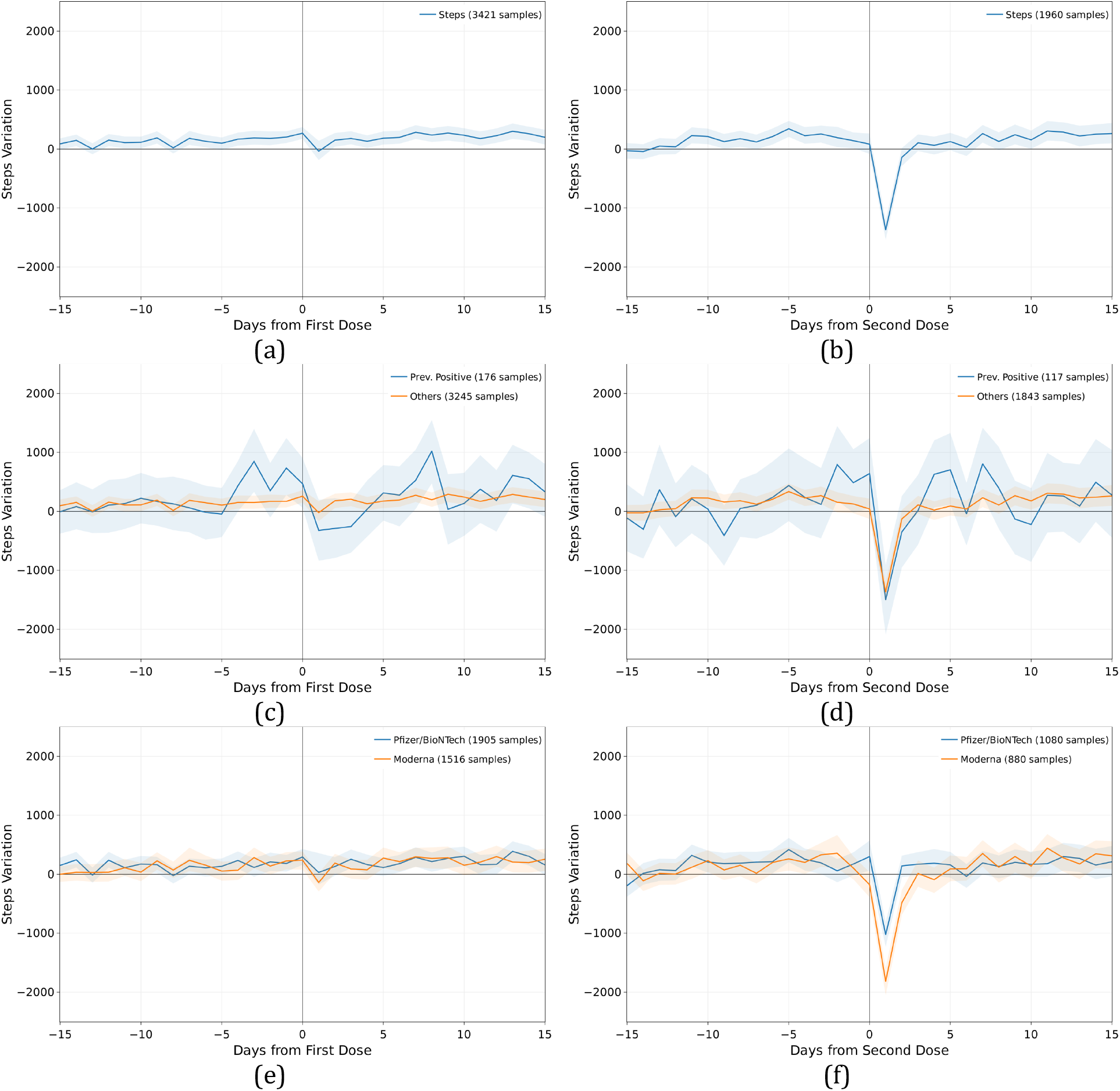
Mean and 95% confidence interval of the absolute individual changes in activity metric (number of steps) with respect to the individual baseline around the date of vaccination (day 0), for the first dose of the vaccine (a), (c), (e), and for the second dose (b), (d), (f), for all individuals vaccinated (a) and (b), for individuals previously tested positive to COVID-19 (c) and (d), and for individuals vaccinated with the Pfizer-BioNTech or Moderna vaccines (e) and (f).

## DISCUSSION

While there is a goal of achieving COVID-19 immunization in billions of people, there is currently no objective means of detecting a response to vaccines except for the occurrence of side-effects. Moreover, while vaccines against COVID-19 are remarkably effective, they are not 100% effective, as recently highlighted by reports of “vaccine breakthrough” in fully vaccinated people. Many individuals are concerned about their lack of any symptoms after getting vaccinated. The present study demonstrates the ability, through widely available wearable sensors, to recognize individual physiologic changes associated with a systemic vaccine-induced inflammatory response.

Individual response to vaccination is remarkably complex, incorporating components of innate, humoral and cell-based immune system. A study of response to yellow fever vaccination found significant modulation of expression in 97 genes in the days following vaccination.^16^ Modern improvements in a range of analytic tools have enabled a system biological approach to better understand the immune responses to vaccination, which have shown promise in helping define molecular signatures that may predict vaccine-induced immunity.^17^ There are no commercially available tests for neutralizing antibodies to the spike protein or its components S1, S2, RBD, that would provide quantitative evidence of an immune response. Beyond humoral immunity, the early T-cell spike-specific response has recently been shown to be important,^18^ yet is only rarely assessed in academic research settings. Accordingly, it is currently impossible to identify, at scale, the level of protection an individual acquires after vaccination.^19^

Currently available mRNA vaccines (Pfizer-BioNTech and Moderna) and adenovirus vaccine (Janssen) elicit an inflammatory response through immune cell activation leading to the production of Type 1 interferon and the release of multiple inflammatory mediators.^20^ Vaccination has been shown to stimulate the production of neutralizing antibodies, activate virus-specific CD4+ and CD8+ T cells, and lead to the robust release of immune-modulatory cytokines in the days that follow a first, and especially a second dose of the mRNA vaccines.^21^ Beyond rapid stimulation of innate immunity via adjuvant stimulation, prior studies of mRNA vaccines have shown peak production of the vaccine-induced antigen protein to occur as soon as 6 hours after vaccination, suggesting that an inflammatory response begins within hours of vaccination.^22^ Consistent with that, we identified a rapid rise in heart rate the day after vaccination, and one that was more robust after the second dose, unless the participant had prior COVID-19 infection, mirroring the significantly higher incidence of systemic symptoms following the second dose found in V-safe.^23^ We also observed a more pronounced increase after a Moderna vaccine, in accordance to a recent analysis of V-safe data that identified a higher incidence of side effects relative to those receiving the Pfizer-BioNTech vaccine, especially after the second dose.^12^ This may be related to a possible unnecessary excess dose and consequent side-effects, supporting the finding of a randomized trial of full (100 ml) versus half (50 ml) dose of the Moderna vaccine with lack of differences in immune response.^24^

Immunosenescence or waning response to vaccination as someone ages has been described for many vaccines and is a concern for COVID-19.^25^ We found that individuals in the younger age group (<40 years) had a significantly higher RHR response to the second dose compared to older individuals. Overall, women also reported more side effects to V-Safe compared to men.^23^ Immune response to other vaccines has varied by gender, possibly because of differences in hormones, genetics, or differences in dosing by weight. A prior flu vaccine study found that vaccine induced immunity in mice was increased by estradiol in females and decreased by testosterone in males^26^ and that as age increased, sex differences in vaccine efficacy was declined. Although the RHR differences we saw was not significant, it is possible that a greater change would be identified when younger age groups are vaccinated.

We found that the observable variables (age, gender, previously COVID-19 infection, device used, vaccine type) can explain only 2% of the variance in terms of average changes in RHR (and less than 24% of the variance in terms of peak changes in RHR). It is possible that with further investigation it may be found that RHR response to vaccines may correlate with individual immune response and therefore wearables may offer a way to easily quantify someone’s protection.

The presence of a fever has previously been shown to be associated with an increase in heart rate, with an ∼8.5 BPM increase for every degree Celsius increase in body temperature.^27^ While ∼30% of V-safe participants reported having a fever after their second dose and ∼9% after the first, we showed that the vast majority of participants experienced an increase in RHR after both vaccination doses, suggesting that inflammation unassociated with an elevated temperature also influences heart rate, albeit much more subtly.^12^ Inflammation, outside of that associated with fever, and its influence on heart rate has been previously described, but primarily at a population level.^28,29^ Similarly, inflammation has also been shown to lead to increase in sleep and a decrease in activity, with a more rapid return to normal with treatment.^30,31^ By taking advantage of wearable sensors we were able to recognize subtle, but significant deviations from an individual’s unique, normal resting heart rate due to vaccination. We were able to also demonstrate substantial interindividual variability in that heart rate increase that was only related to the mRNA vaccine type and prior COVID-19 infection in our population. The significantly greater heart rate response at the time of vaccination, especially the first dose, in those with prior infection is consistent with a greater immune response for these individuals.^32^ Although this response is clearly multifactorial, future work coupling continuous sensory data with a “systems vaccinology” approach^17^ could enable the identification of easily scalable, sensor-based markers of the desired immune response.

### Limitations

The data collected as part of the DETECT study depends entirely on the participants’ willingness to use their wearable device and accurately reporting vaccination date and type. While we do not have direct control on self-reported information, the DETECT app provides an intuitive tool to self-report vaccination information, and an optional reminder to report information on the second dose after first one has been reported. While the information collected may not be as accurate as in a controlled laboratory setting, we rely on previous work confirming that self-reported symptoms and sensor data provide valuable information.^33-35^ Only daily sensor data is considered in this analysis, excluding intra-day data provided by some wearable sensor. These once-a-day values are indeed more stable and less affected by independent confounders like the specific activity performed by the individual during the day. Furthermore, the population in the DETECT study that has received a COVID-19 vaccine may not be representative of the population of the United States, as the study is open to individuals who have access to a wearable device technology.^36^ While research has found no racial or ethnic variation in smartwatch or activity tracker usage in the U.S., they are less commonly utilized by older individuals, those in the lowest socioeconomic tertile, and lower educational attainment.^37^ Another reason our results may not be representative of the broader population is that the first phase of the vaccine distribution has focused on an older age range and at-risk individuals based on their profession or health status. It is also possible that some participants had prior COVID-19 that went undiagnosed, which may have impacted their immune response to the first dose.

## Conclusions

Fitness bands and smartwatches are owned by approximately 1 in 3 American adults and appear to provide, through passive data capture, meaningful data to track the physiologic response to COVID-19 vaccines at the individual level. Not only might this provide reassurance for vaccinees who do not experience any symptoms, but correlation with the humoral and cellular immune response may indicate digital tracking as a useful surrogate. Noteworthy is the potential to identify the small proportion of people (approximately 5%) who do not have an adequate immune response to vaccines, and who may benefit by more in-depth assessment and re-vaccination.

## Data Availability

This is an ongoing study. We plan to make the de-identified data available after approval of a proposal by a responsible authority at Scripps and with a data access agreement, pledging to not re-identify individuals or share the data with a third party.

## ACKNOWLEDGEMENTS

This work was funded by grant number UL1TR002550 from the National Center for Advancing Translational Sciences (NCATS) at the National Institutes of Health (NIH) (E.J.T., K.G.A, and S.R.S.) and NIH NIAID grant number U19AI135995 (K.G.A and S.R.S). We thank also Danny Oran, Lauren Ariniello, Tyler Peters, Royan Kamyar, Chris Nowak and Vik Kheterpal for their contribution to DETECT.

## AUTHORS CONTRIBUTIONS

G.Q., M.G., and S.R.S. made substantial contributions to the study conception and design. G.Q., M.G., J.M.R., K.B.-M., E.R., and S.R.S. made substantial contributions to the acquisition of data. G.Q., M.G., and J.M.R. conducted statistical analysis. G.Q., M.G., J.M.R., and S.R.S. made substantial contributions to the interpretation of data. G.Q., M.G., and S.R.S. drafted the first version of the manuscript. G.Q., M.G., J.M.R., K.G.A., K.B.-M., E.R., E.J.T. and S.R.S. contributed to critical revisions and approved the final version of the manuscript. G.Q., M.G. and S.R.S. take responsibility for the integrity of the work.

## DECLARATION OF INTERESTS

S.R.S. is employed by PhysIQ. The other authors declare no competing interests.

## SUPPLEMENTAL TABLES

**Table S.1.** Demographic characteristics (age and gender) and device used by individuals in the 4 groups used in the analysis, depending on vaccine type and if previously tested positive to COVID-19. A chi-squared test has been used to evaluate significant changes in the frequency of observation in each group.

|  |  | After First Dose |  |  |  |  |  | After Second Dose |  |  |  |  |  |
| --- | --- | --- | --- | --- | --- | --- | --- | --- | --- | --- | --- | --- | --- |
|  |  | Individuals | Age<br>Median<br>[IQR] | Female | Age >60 | Prev.<br>Positive | Fitbit | Individuals | Age<br>Median<br>[IQR] | Female | Age >60 | Prev.<br>Positive | Fitbit |
| RHR Variation<br>[BPM] | Overall | 3312 | 57<br>[45 - 66] | 57.20% | 40.70% | 5.00% | 74.90% | 1905 | 61<br>[47 - 69] | 58.70% | 50.20% | 5.70% | 73.30% |
|  | Moderna | 1465 | 58<br>[46 - 67] | 58.2%<br>p=0.32 | 43.8%<br>p < 0.01 | 4.3%<br>p=0.13 | 75.3%<br>p=0.71 | 865 | 62<br>[48 - 70] | 60.0%<br>p=0.33 | 52.3%<br>p=0.12 | 6.2%<br>p=0.37 | 73.8%<br>p=0.74 |
|  | Pfizer/BioNTech | 1847 | 56<br>[44 - 65] | 56.4%<br>p=0.32 | 38.2%<br>p < 0.01 | 5.5%<br>p=0.13 | 74.7%<br>p=0.71 | 1040 | 60<br>[47 - 68] | 57.7%<br>p=0.33 | 48.6%<br>p=0.12 | 5.2%<br>p=0.37 | 73.0%<br>p=0.74 |
|  | Prev. Positive | 165 | 52<br>[44 - 62] | 67.9%<br>p < 0.01 | 30.3%<br>p < 0.01 | - | 78.2%<br>p=0.37 | 108 | 53<br>[44 - 66] | 75.0%<br>p < 0.01 | 36.1%<br>p < 0.01 | - | 79.6%<br>p=0.16 |
|  | Others | 3147 | 57<br>[45 - 66] | 56.6%<br>p < 0.01 | 41.2%<br>p < 0.01 | - | 74.8%<br>p=0.37 | 1797 | 61<br>[48 - 69] | 57.8%<br>p < 0.01 | 51.1%<br>p < 0.01 | - | 73.0%<br>p=0.16 |
| Sleep Variation<br>[min] | Overall | 2675 | 57<br>[44 - 66] | 57.80% | 39.80% | 5.30% | 86.20% | 1490 | 60<br>[47 - 69] | 59.40% | 49.90% | 5.90% | 85.50% |
|  | Moderna | 1174 | 58<br>[45 - 67] | 58.9%<br>p=0.29 | 42.2%<br>p=0.02 | 4.4%<br>p=0.10 | 86.7%<br>p=0.51 | 671 | 61<br>[47 - 69] | 61.0%<br>p=0.29 | 51.3%<br>p=0.35 | 6.4%<br>p=0.53 | 86.6%<br>p=0.32 |
|  | Pfizer/BioNTech | 1501 | 56<br>[44 - 65] | 56.8%<br>p=0.29 | 37.8%<br>p=0.02 | 5.9%<br>p=0.10 | 85.7%<br>p=0.51 | 819 | 60<br>[47 - 68] | 58.1%<br>p=0.29 | 48.7%<br>p=0.35 | 5.5%<br>p=0.53 | 84.6%<br>p=0.32 |
|  | Prev. Positive | 141 | 52<br>[43 - 62] | 67.4%<br>p=0.02 | 30.5%<br>p=0.03 | - | 87.2%<br>p=0.80 | 88 | 51<br>[42 - 65] | 72.7%<br>p=0.01 | 36.4%<br>p=0.01 | - | 92.0%<br>p=0.10 |
|  | Others | 2534 | 57<br>[45 - 66] | 57.2%<br>p=0.02 | 40.3%<br>p=0.03 | - | 86.1%<br>p=0.80 | 1402 | 61<br>[47 - 69] | 58.6%<br>p=0.01 | 50.7%<br>p=0.01 | - | 85.1%<br>p=0.10 |
| Steps Variation | Overall | 3421 | 57<br>[45 - 66] | 57.60% | 40.20% | 5.10% | 77.50% | 1960 | 60<br>[47 - 69] | 59.20% | 49.40% | 6.00% | 76.50% |
|  | Moderna | 1516 | 58<br>[46 - 67] | 58.7%<br>p=0.26 | 43.1%<br>p < 0.01 | 4.4%<br>p=0.10 | 77.5%<br>p=0.98 | 880 | 62<br>[48 - 69] | 60.5%<br>p=0.32 | 51.7%<br>p=0.08 | 6.5%<br>p=0.45 | 76.8%<br>p=0.79 |
|  | Pfizer/BioNTech | 1905 | 56<br>[44 - 65] | 56.7%<br>p=0.26 | 37.8%<br>p < 0.01 | 5.7%<br>p=0.10 | 77.5%<br>p=0.98 | 1080 | 59<br>[47 - 68] | 58.1%<br>p=0.32 | 47.6%<br>p=0.08 | 5.6%<br>p=0.45 | 76.2%<br>p=0.79 |
|  | Prev. Positive | 176 | 52<br>[44 - 62] | 69.3%<br>p < 0.01 | 29.5%<br>p < 0.01 | - | 79.5%<br>p=0.57 | 117 | 53<br>[42 - 64] | 74.4%<br>p < 0.01 | 34.2%<br>p < 0.01 | - | 81.2%<br>p=0.26 |
|  | Others | 3245 | 57<br>[45 - 66] | 57.0%<br>p < 0.01 | 40.8%<br>p < 0.01 | - | 77.4%<br>p=0.57 | 1843 | 61<br>[47 - 69] | 58.2%<br>p < 0.01 | 50.4%<br>p < 0.01 | - | 76.2%<br>p=0.26 |

**Table S.2.** Coefficients with 95% confidence interval and associated significance for a multiple regression model to predict the average RHR change after each dose of one mRNA vaccine. Age has been normalized with respect to the population median.

|  | First Dose |  |  | Second Dose |  |  |
| --- | --- | --- | --- | --- | --- | --- |
|  | Coefficient | Std. Error | p-value | Coefficient | Std. Error | p-value |
| **(Intercept)** | 0.253 | 0.180 | 0.160 | 1.395 | 0.237 | <0.01 |
| **Normalized Age** | -0.066 | 0.176 | 0.708 | -0.923 | 0.224 | <0.01 |
| **Gender (1 if Male)** | -0.057 | 0.086 | 0.509 | -0.068 | 0.113 | 0.547 |
| **Prev. Covid (1 if Positive)** | 0.477 | 0.194 | 0.014 | -0.491 | 0.237 | 0.039 |
| **VaccineType (1 if Moderna)** | 0.180 | 0.085 | 0.035 | 0.449 | 0.110 | <0.01 |
| **Device (1 if Apple)** | 0.091 | 0.098 | 0.352 | 0.172 | 0.124 | 0.165 |

## Notes

### Author Declarations

The protocol for DETECT was reviewed and approved by the Scripps Office for the Protection of Research Subjects (IRB 20-7531). All participants in the study provided informed consent electronically.

## REFERENCES

1. Oliver, S.E., et al. The Advisory Committee on Immunization Practices’ Interim Recommendation for Use of Pfizer-BioNTech COVID-19 Vaccine - United States, December 2020. MMWR Morb Mortal Wkly Rep 69, 1922–1924 (2020).

2. Oliver, S.E., et al. The Advisory Committee on Immunization Practices’ Interim Recommendation for Use of Moderna COVID-19 Vaccine - United States, December 2020. MMWR Morb Mortal Wkly Rep 69, 1653–1656 (2021).

3. Oliver, S.E., et al. The Advisory Committee on Immunization Practices’ Interim Recommendation for Use of Janssen COVID-19 Vaccine - United States, February 2021. MMWR Morb Mortal Wkly Rep 70, 329–332 (2021).

4. Polack, F.P., et al. Safety and Efficacy of the BNT162b2 mRNA Covid-19 Vaccine. N Engl J Med 383, 2603–2615 (2020).

5. Aran, D. Estimating real-world COVID-19 vaccine effectiveness in Israel using aggregated counts. medRxiv, 2021.2002.2005.21251139 (2021).

6. Baden, L.R., et al. Efficacy and Safety of the mRNA-1273 SARS-CoV-2 Vaccine. N Engl J Med 384, 403–416 (2021).

7. Sadoff, J., et al. Interim Results of a Phase 1-2a Trial of Ad26.COV2.S Covid-19 Vaccine. N Engl J Med (2021).

8. Thompson, M.G., et al. Interim Estimates of Vaccine Effectiveness of BNT162b2 and mRNA-1273 COVID-19 Vaccines in Preventing SARS-CoV-2 Infection Among Health Care Personnel, First Responders, and Other Essential and Frontline Workers — Eight U.S. Locations. MMWR Morb Mortal Wkly Rep ePub: (2021).

9. Zimmermann, P. & Curtis, N. Factors That Influence the Immune Response to Vaccination. Clin Microbiol Rev 32(2019).

10. Poland, G.A. & Jacobson, R.M. Failure to Reach the Goal of Measles Elimination: Apparent Paradox of Measles Infections in Immunized Persons. Archives of Internal Medicine 154, 1815–1820 (1994).

11. NIH. V-safe After Vaccination Health Checker. https://www.cdc.gov/coronavirus/2019-ncov/vaccines/safety/vsafe.html. (2021).

12. Chapin-Bardales, J., Gee, J.& Myers, T. Reactogenicity Following Receipt of mRNA-Based COVID-19 Vaccines. Jama (2021).

13. Quer, G., et al. Wearable sensor data and self-reported symptoms for COVID-19 detection. Nature Medicine 27, 73–77 (2021).

14. Quer, G., Gouda, P., Galarnyk, M.,Topol, E.J. & Steinhubl, S.R. Inter-and intraindividual variability in daily resting heart rate and its associations with age, sex, sleep, BMI, and time of year: Retrospective, longitudinal cohort study of 92,457 adults. PLoS One 15, e0227709 (2020).

15. Jaiswal, S.J., et al. Association of Sleep Duration and Variability With Body Mass Index: Sleep Measurements in a Large US Population of Wearable Sensor Users. JAMA Internal Medicine (2020).

16. Querec, T.D., et al. Systems biology approach predicts immunogenicity of the yellow fever vaccine in humans. Nat Immunol 10, 116–125 (2009).

17. Nakaya, H.I. & Pulendran, B. Vaccinology in the era of high-throughput biology. Philos Trans R Soc Lond B Biol Sci 370(2015).

18. Kalimuddin, S., et al. Early T cell and binding antibody responses are associated with Covid-19 RNA vaccine efficacy onset. Med (2021).

19. Brodin, P. & Davis, M.M. Human immune system variation. Nat Rev Immunol 17, 21–29 (2017).

20. Teijaro, J.R. & Farber, D.L. COVID-19 vaccines: modes of immune activation and future challengess. Nature Reviews Immunology (2021).

21. Sahin, U., et al. COVID-19 vaccine BNT162b1 elicits human antibody and TH1 T cell responses. Nature 586, 594–599 (2020).

22. Bahl, K., et al. Preclinical and Clinical Demonstration of Immunogenicity by mRNA Vaccines against H10N8 and H7N9 Influenza Viruses. Mol Ther 25, 1316–1327 (2017).

23. Gee, J., et al. First Month of COVID-19 Vaccine Safety Monitoring - United States, December 14, 2020-January 13, 2021. MMWR Morb Mortal Wkly Rep 70, 283–288 (2021).

24. Chu, L., et al. A preliminary report of a randomized controlled phase 2 trial of the safety and immunogenicity of mRNA-1273 SARS-CoV-2 vaccine. Vaccine (2021).

25. Cox, L.S., et al. Tackling immunosenescence to improve COVID-19 outcomes and vaccine response in older adults. Lancet Healthy Longev 1, e55–e57 (2020).

26. Potluri, T., et al. Age-associated changes in the impact of sex steroids on influenza vaccine responses in males and females. npj Vaccines 4, 29 (2019).

27. Karjalainen, J. & Viitasalo, M. Fever and cardiac rhythm. Arch Intern Med 146, 1169–1171 (1986).

28. Whelton, S.P., et al. Association between resting heart rate and inflammatory biomarkers (high-sensitivity C-reactive protein, interleukin-6, and fibrinogen) (from the Multi-Ethnic Study of Atherosclerosis). Am J Cardiol 113, 644–649 (2014).

29. Park, W.C., Seo, I., Kim, S.H., Lee, Y.J. & Ahn, S.V. Association between Resting Heart Rate and Inflammatory Markers (White Blood Cell Count and High-Sensitivity CReactive Protein) in Healthy Korean People. Korean J Fam Med 38, 8–13 (2017).

30. Bryant, P.A., Trinder, J. & Curtis, N. Sick and tired: does sleep have a vital role in the immune system? Nature Reviews Immunology 4, 457–467 (2004).

31. Bettis, R., et al. Impact of influenza treatment with oseltamivir on health, sleep and daily activities of otherwise healthy adults and adolescents. Clin Drug Investig 26, 329–340 (2006).

32. Ebinger, J.E., et al. Antibody responses to the BNT162b2 mRNA vaccine in individuals previously infected with SARS-CoV-2. Nature Medicine (2021).

33. Rivera, S.C., et al. The impact of patient-reported outcome (PRO) data from clinical trials: a systematic review and critical analysis. Health and Quality of Life Outcomes 17, 156 (2019).

34. Bell, S.K., et al. Frequency and Types of Patient-Reported Errors in Electronic Health Record Ambulatory Care Notes. JAMA Network Open 3, e205867–e205867 (2020).

35. Basch, E., et al. Overall Survival Results of a Trial Assessing Patient-Reported Outcomes for Symptom Monitoring During Routine Cancer Treatment. Jama 318, 197–198 (2017).

36. Beaunoyer, E., Dupéré, S. & Guitton, M.J. COVID-19 and digital inequalities: Reciprocal impacts and mitigation strategies. Comput Human Behav 111, 106424 (2020).

37. Vogels, E.A. About one-in-five Americans use a smart watch or fitness tracker, https://www.pewresearch.org/fact-tank/2020/01/09/about-one-in-five-americans-use-a-smart-watch-or-fitness-tracker/. (2020).

